# Gender differences in patients with COVID-19: Focus on severity and mortality

**DOI:** 10.1101/2020.02.23.20026864

**Authors:** Jian-Min Jin, Peng Bai, Wei He, Fei Wu, Xiao-Fang Liu, De-Min Han, Shi Liu, Jin-Kui Yang

## Abstract

**Importance:** The recent outbreak of Novel Coronavirus (SARS-CoV-2) Disease (COVID-19) has put the world on alert, that is reminiscent of the SARS outbreak seventeen years ago.

**Objective:** We aim to compare the severity and mortality between male and female patients with both COVID-19 and SARS, to explore the most useful prognostic factors for individualized assessment.

**Design, Setting, and Participants:** We extracted the data from a case series of 43 hospitalized patients we treated, a public data set of the first 37 cases died of COVID-19 in Wuhan city and 1019 survived patients from six cities in China. We also analyzed the data of 524 patients with SARS, including 139 deaths, from Beijing city in early 2003.

**Main Outcomes and Measures:** Severity and mortality.

**Results:** Older age and high number of comorbidities were associated with higher severity and mortality in patients with both COVID-19 and SARS. The percentages of older age (≥65 years) were much higher in the deceased group than in the survived group in patients with both COVID-19 (83.8 vs. 13.2, P<0.001) and SARS (37.4 vs. 4.9, P<0.001). In the case series, men tend to be more serious than women (P=0.035), although age was comparable between men and women. In the public data set, age was also comparable between men and women in the deceased group or the survived group in patients with COVID-19. Meanwhile, gender distribution was exactly symmetrical in the 1019 survivors of COVID-19. However, the percentage of male were higher in the deceased group than in the survived group (70.3 vs. 50.0, P=0.015). The gender role in mortality was also observed in SARS patients. Survival analysis showed that men (hazard ratio [95% CI] 1.47 [1.05-2.06, P= 0.025) had a significantly higher mortality rate than women in patients with SARS.

**Conclusions and Relevance:** Older age and male gender are risk factors for worse outcome in patients with COVID. While men and women have the same susceptibility to both SARS-CoV-2 and SARS-CoV, men may be more prone to have higher severity and mortality independent of age and susceptibility.

**Key Points:** *Question:* Are men more susceptible to getting and dying from COVID-19?

*Findings:* In the case series, men tend to be more serious than women. In the public data set, the percentage of men were higher in the deceased group than in the survived group, although age was comparable between men and women.

*Meaning:* Male gender is a risk factor for worse outcome in patients with COVID independent of age and susceptibility.

## Introduction

In early December 2019, an outbreak of a novel coronavirus disease (COVID-19) in Wuhan city and the rapidly spread throughout China has put the world on alert. High-throughput sequencing has revealed a novel β-coronavirus that is currently named as severe acute respiratory syndrome coronavirus 2 (SARS-CoV-2)^1^, which resembled severe acute respiratory syndrome coronavirus (SARS-CoV)^2^. Most patients with COVID-19 were mild. Moderate patients often experienced dyspnea after one week. Severe patients progressed rapidly to critical conditions including acute respiratory distress syndrome (ARDS), acute respiratory failure, coagulopathy, septic shock, and metabolic acidosis.

Early identification of risk factors for critical conditions is urgently warranted not only to identify the defining clinical and epidemiological characteristics with greater precision, but also facilitated appropriate supportive care and promptly access to the intensive care unit (ICU) if necessary.

The Chinese health authority has announced that the total number of confirmed cases on the Chinese mainland has reached 76,936, and 2,442 people have died of the disease as of Feb 23. Among the 2,442 deceased patients, most were old and two-thirds were males, though the detailed data has not been reported^3^. This raises a question: Are men more susceptible to getting and dying from COVID-19?

Here, we reported the clinical characteristics of a recent case series of 43 patients with 2019 SARS-CoV-2 infection we treated, a public data set of the first 37 cases died of SARS-CoV-2 in Wuhan city and the 1019 survived patients with SARS-CoV-2 from six cities with a high prevalence of the disease. We aimed to compare the severity and mortality in male and female patients with COVID-19, to explore the most useful prognostic factor for individualized assessment. SARS-CoV-2 infection is reminiscent of the SARS-CoV outbreak in early 2003, because both viruses attack cells via the same ACE2 receptor^3^. In this study, we also analyzed the data of 524 SARS patients, including 139 deaths, from Beijing in early 2003.

## Methods

### Patients and data sources

In the case series analysis, a recent case series of 43 patients with COVID-19 was treated at Wuhan Union Hospital by the medical team of Beijing Tongren Hospital from January 29, 2020 to February 15, 2020. Respiratory specimens were collected by local center for disease control and prevention (CDC) and then shipped to designated authoritative laboratories to detect SARS-CoV-2. The presence of SARS-CoV-2 in respiratory specimens was detected by real-time RT-PCR methods. The RT-PCR assay was conducted in accordance with the protocol established by the World Health Organization.

In the public data set, the first 37 cases died of COVID-19 in Wuhan city and the 1019 survived patients with COVID-19 from six cities with a high prevalence of the disease were obtained from the Chinese Public Health Science Data Center. Case data included basic demographic information, classification, date of symptom onset, date of diagnosis, date of hospitalization, date of discharge or date of death, etc.

Diagnosis and clinical classification criteria and treatment plan (trial version 5) of the SARS-CoV-2 coronavirus pneumonia (COVID-19) was launched by the National Health Committee of the People’s Republic of China (http://www.nhc.gov.cn/). The clinical classification of severity is as follows: (1) Mild, only mild symptoms, imaging shows no pneumonia. (2) Common, with fever, respiratory tract symptoms, and imaging shows pneumonia. (3) Severe, meet any of the following signs: a) respiratory distress, respiratory rate ≥ 30 beats / min; b) in the resting state, finger oxygen saturation ≤ 93%) arterial blood oxygen partial pressure (PaO_2_/oxygen concentration (FiO_2_) ≤ 300mmHg (1mmHg = 0.133kPa). (4) Critical, one of the following conditions: a) respiratory failure occurs and requires mechanical ventilation; b) Shock occurs; c) ICU admission is required for combined organ failure.

This study also included data of 524 SARS patients, including 139 deaths, from Beijing in early 2003. These patients were hospitalized in Beijing between 25 March and 22 May 2003.

The study protocol was approved by the Ethics Committee of Beijing Tongren Hospital, Capital Medical University.

### Statistical analysis

Data were expressed as mean ± SD, median (interquartile range (IQR)) or percentage, as appropriate. Compared the differences between the two groups, mean values and percentages were compared between the two groups by the Student *t*-test, Mann-Whitney U test or chi-square (χ^2^) test. Kaplan–Meier survival curves and the log-rank test was used for testing the survival between males and females. Statistical analyses were performed using the SAS software (version 9.4). *P*< 0.05 (two-tailed) was considered to be statistically significant.

## Results

### Case series of COVID-19 we treated

The demographic and clinical characteristics are shown in Table 1. The median age was 62 years (IQR, 51 to 70). Fever (95.3%) and cough (65.1%) were the most common symptoms, while diarrhea (16.3) was not common. 37.2% of patients had at least one underlying disorder (i.e., hypertension, diabetes, cardiovascular diseases and chronic lung diseases). Ages, symptoms and comorbidities were comparable between men and women. As expected, men had a higher level of hemoglobin. However, male patients also had elevated serum creatinine, white blood cells and neutrophils. Among the 43-case series, 13 (30.2%) were diagnosed with mild or common pneumonia, 14 (32.6%) and 16 (37.2%) were diagnosed with severe and critical pneumonia, respectively. Chi-square (χ^2^) test for trend indicated that men tend to be more serious than women (*P*=0.035) according to the clinical classification of severity (Figure 1).

**Table 1.** Characteristics of a case series of 43 patients with COVID.

|  | Total<br>(n=43) | Male<br>(n=22) | Female<br>(n=21) | P-value |
| --- | --- | --- | --- | --- |
| Age, median (range) – year | 62 (51-70) | 59 (51-66) | 63 (52-73) | 0.734 |
| Symptoms |  |  |  |  |
| Fever – n (%) | 41 (95.3) | 21 (95.5) | 20 (95.2) | 0.490 |
| Diarrhea –n (%) | 7 (16.3) | 3 (13.6) | 4 (19.0) | 0.946 |
| Cough – n (%) | 28 (65.1) | 16 (72.7) | 12 (57.1) | 0.452 |
| Comorbidities –n (%) |  |  |  |  |
| Hypertension –n (%) | 10 (23.3) | 6 (27.3) | 4 (19.0) | 0.782 |
| Diabetes history –n (%) | 5 (11.6) | 4 (18.2) | 1 (0.5) | 0.370 |
| Cardiovascular diseases –n (%) | 4 (9.3) | 2 (9.1) | 2 (10.0) | 0.634 |
| Chronic lung diseases –n (%) | 1 (0.2) | 1 (0.5) | 0 (0) | 0.981 |
| From symptom to diagnosis, median (range) – day | 12 (8-14) | 12 (7-13) | 12 (10-14) | 0.250 |
| Aspartate aminotransferase – IU/l | 42.4±18.9 | 43.0±15.3 | 41.7±22.6 | 0.872 |
| Alanine aminotransferase – IU/l | 42.8±19.0 | 45.0±18.0 | 40.4±19.5 | 0.590 |
| Alkaline phosphatase – IU/l | 53.4±10.6 | 52.6±11.9 | 54.3±9.0 | 0.736 |
| Lactate dehydrogenase – IU/l | 369.4±132.7 | 414.8±136.2 | 321.8±112.9 | 0.064 |
| Serum creatinine – µmol/l | **75.3±21.1** | **90.4±22.2** | **59.4±10.9** | **0.000** |
| Fasting Blood Glucose – mmol/l | 7.3±1.8 | 7.7±2.0 | 6.7±1.5 | 0.325 |
| High sensitive C-reactive protein – mg/l | 52.3±27.8 | 58.9±29.2 | 45.6±25.3 | 0.323 |
| White blood cells – ×10<sup>9</sup>/l | **6.8±2.2** | **7.7±2.3** | **5.8±1.5** | **0.027** |
| Hemoglobin – g/l | 128.8±13.6 | 139.0±11.2 | 117.6±8.6 | 0.000 |
| Platelets – ×10<sup>9</sup>/l | 225.2±57.4 | 230.4±54.1 | 219.6±60.0 | 0.682 |
| Neutrophils – ×10<sup>9</sup>/l | **5.4±2.2** | **6.4±2.4** | **4.3±1.3** | **0.019** |
| Lymphocytes – ×10<sup>9</sup>/l | 1.0±0.4 | 0.9±0.3 | 1.1±0.4 | 0.284 |

Data are presented as mean ± SD, medians (interquartile ranges, IQR) and no. (%).

**Figure 1.**
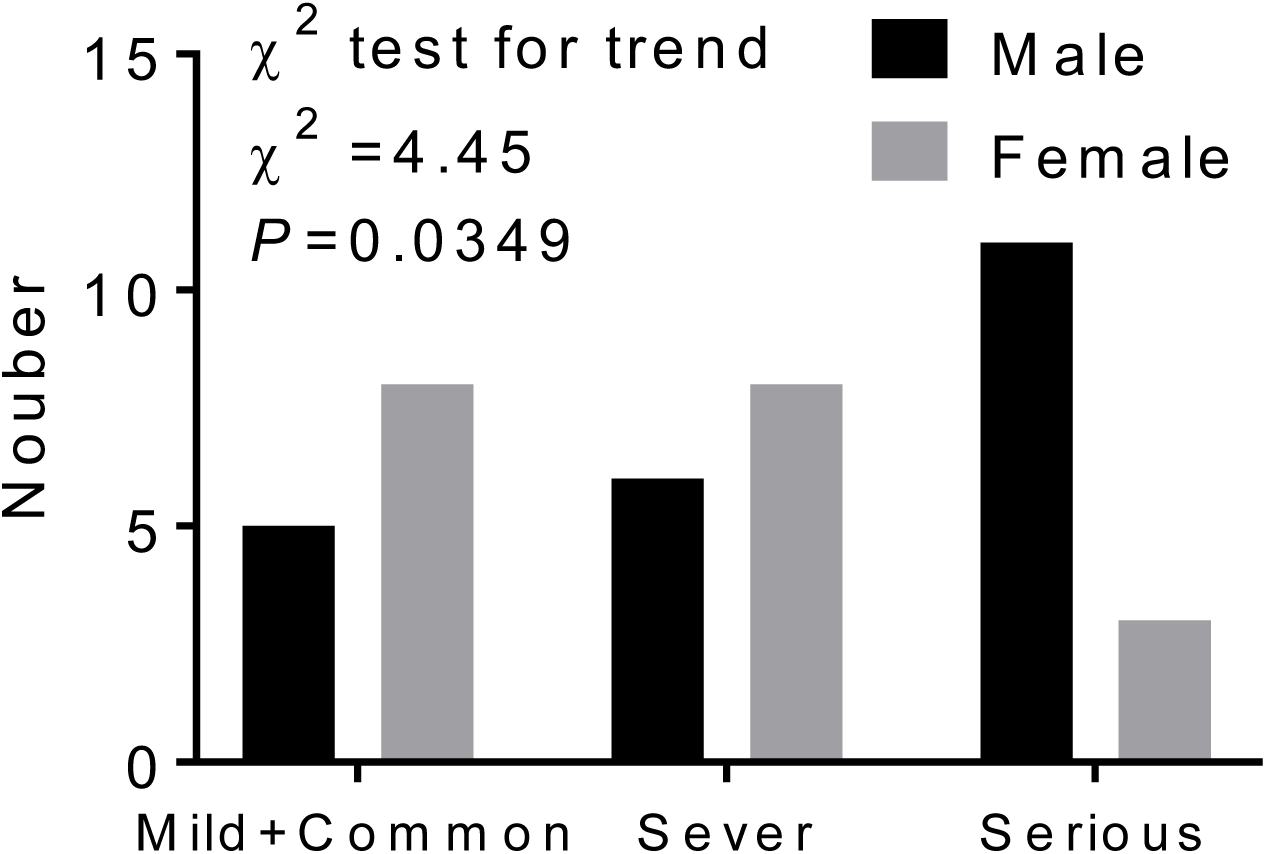
Trend data of clinical classification of severity. Numbers of cases of men or women in different clinical classes of severity. Chi-square (χ^2^) test for trend indicated that males tend to be more serious than females according to the clinical classification of severity including Mild+Common, Severe and Critical.

### Observational study of COVID-19 from the public data set

In the deceased patients, fever (86.5%) and cough (67.6%) were common, while diarrhea was uncommon (18.9%). The median period from symptom onset to death was 13 days (ranging from 11 to 18 days). Of these deceased patients, 64.9% had at least one underlying disorder (i.e., hypertension, diabetes, cardiovascular disease, or chronic obstructive pulmonary disease) (Table 2).

**Table 2.** Characteristics of Patients with COVID-19 and SARS-2003.

|  | COVID-19 |  | SARS |  |
| --- | --- | --- | --- | --- |
|  | Deceased<br>(n=37) | Survived<br>(n=1019) | Deceased<br>(n=139) | Survived<br>(n=385) |
| Age, median (range) – year | 70 (65-81) ** | 47 (35-57) | 57 (45-69) †† | 32 (24-44) |
| Male – n (%) | 26 (70.3)* | 510 (50.0) | 74 (53.2)† | 163 (42.3) |
| Symptoms |  |  |  |  |
| Fever – % | 32 (86.5) |  | 136 (97.8) | 379 (98.4) |
| Diarrhea – % | 7 (18.9) |  | 30 (21.6) | 26 (6.8) |
| Cough – % | 25 (67.6) |  | 107 (77.0) | 185 (48.1) |
| Comorbidities – % | 24 (64.9) |  | 79 (56.8)†† | 69 (17.9) |
| Hypertension – % | 18 (48.6) |  | 64 (46.0)†† | 44 (11.4) |
| Diabetes history – % | 11(29.7) |  | 30 (21.6)†† | 15 (3.9) |
| Cardiovascular disease – % | 8 (21.6) |  | 40 (28.8)†† | 23 (6.0) |
| Chronic lung disease – % | 3 (8.1) |  | 5 (3.6) | 6 (1. 6) |
| From onset to death, median (range) – day | 13 (11-18) |  | 15 (10-19) |  |

Data are presented as medians (interquartile ranges, IQR) and n (%).
\* p<0.05, \*\* p<0.01, vs. COVID-19 survived patients.
† p<0.05, †† p<0.01, vs. SARS survived patients.

The deceased patients were significantly older (median (IQR), 70.3 (65-81) years) and had a higher percentage of ≥65 years (83.8%), in comparison to those who survived (47 (35-57) years old and 13.2% ≥65 years). COVID-19 was diagnosed throughout the whole spectrum of age. There were 30 (2.9%) pediatric patients (<14 years) in the survived group. None of the 37 deceased cases was pediatric patient (Table 2 and Figure 2A). Ages were comparable between men and women in both deceased and survived patients (Figure 2B). Of the 1019 survived patients with COVID-19 from six cities with a high prevalence of the disease, the proportions of male were 47.8% (Shenzhen city, n=278), 48.2% (Wenzhou city, n=199), 48.3% (Beijing city, n=174), 48.6% (Changsha city, n=140), 60.0% (Xinyang city, n=125) and 52.4% (Hefei city, n=103) respectively in these cities. In total, 50.0% were men in survived cases. However, of the 37 deceased patients, 26 (70.3%) were men. While men and women had the same susceptibility, men were more prone to dying (χ^2^ test, P=0.016) (Figure 2C).

**Figure 2.**
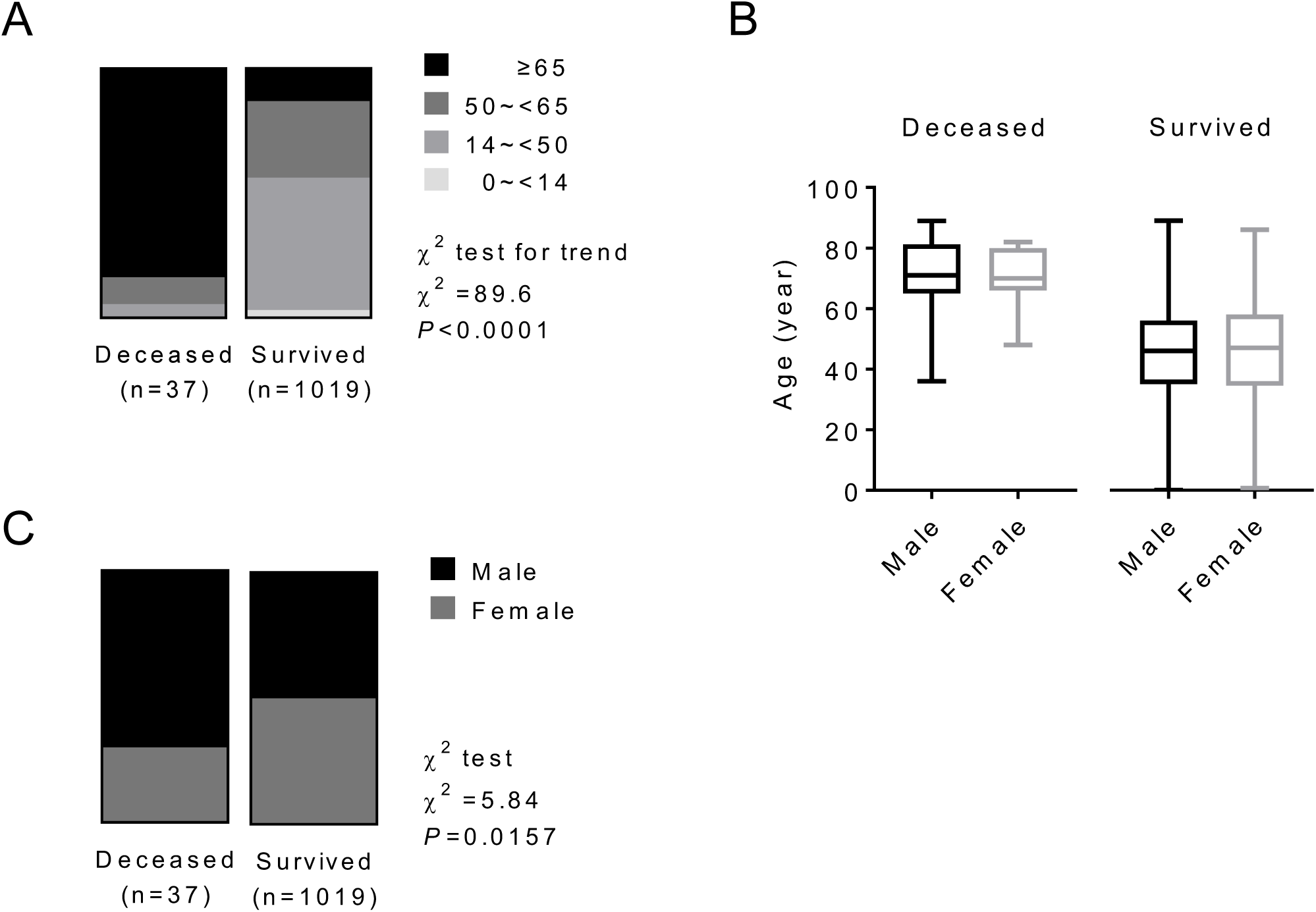
Role of age and gender in morbidity and mortality in patients with COVID-19. **(A)** The whole spectrum of age in deceased and survived patients with COVID. **(B)** Comparation of age between males and females in both deceased and survived patients with COVID. **(C)** Gender distribution in both deceased and survived patients with COVID.

### Observational study of SARS in Beijing, in 2003

Between March 25 and May 22, 2003, a total of 139 patients died of SARS in Beijing area reported from 29 hospitals were enrolled in our analysis. Fever (98.4%) and cough (76.9%) were the most common symptoms, while diarrhea (6.7%) was not common. 57.0% of the patients had at least one of the concomitant diseases including hypertension, diabetes, cardiovascular diseases and chronic lung diseases. The mean duration from self-reported symptom to death was 15 (IQR: 10-19) days. Table 2 summarizes the clinical and biochemical characteristics of all SARS patients. The median age of the deceased patients was much higher than that of the survived patients (57 vs. 32, P<0.001). The rate of the concomitant diseases in the deceased patients was also higher than that of the survived patients (57.0% vs. 17.9%, *P*<0.001). While the deceased patients were significantly older than the survived patients (Figure 3A), ages were comparable between men and women in both deceased and survived patients with SARS (Figure 3B). The proportion of males was higher in the deceased group than in the survived group (χ^2^ test, P=0.016) (Figure 3C). Survival analysis showed that men (hazard ratio [95% CI] 1.47 [1.05-2.06], P= 0.025) had a significantly higher mortality rate than women (Figure 3D).

**Figure 3.**
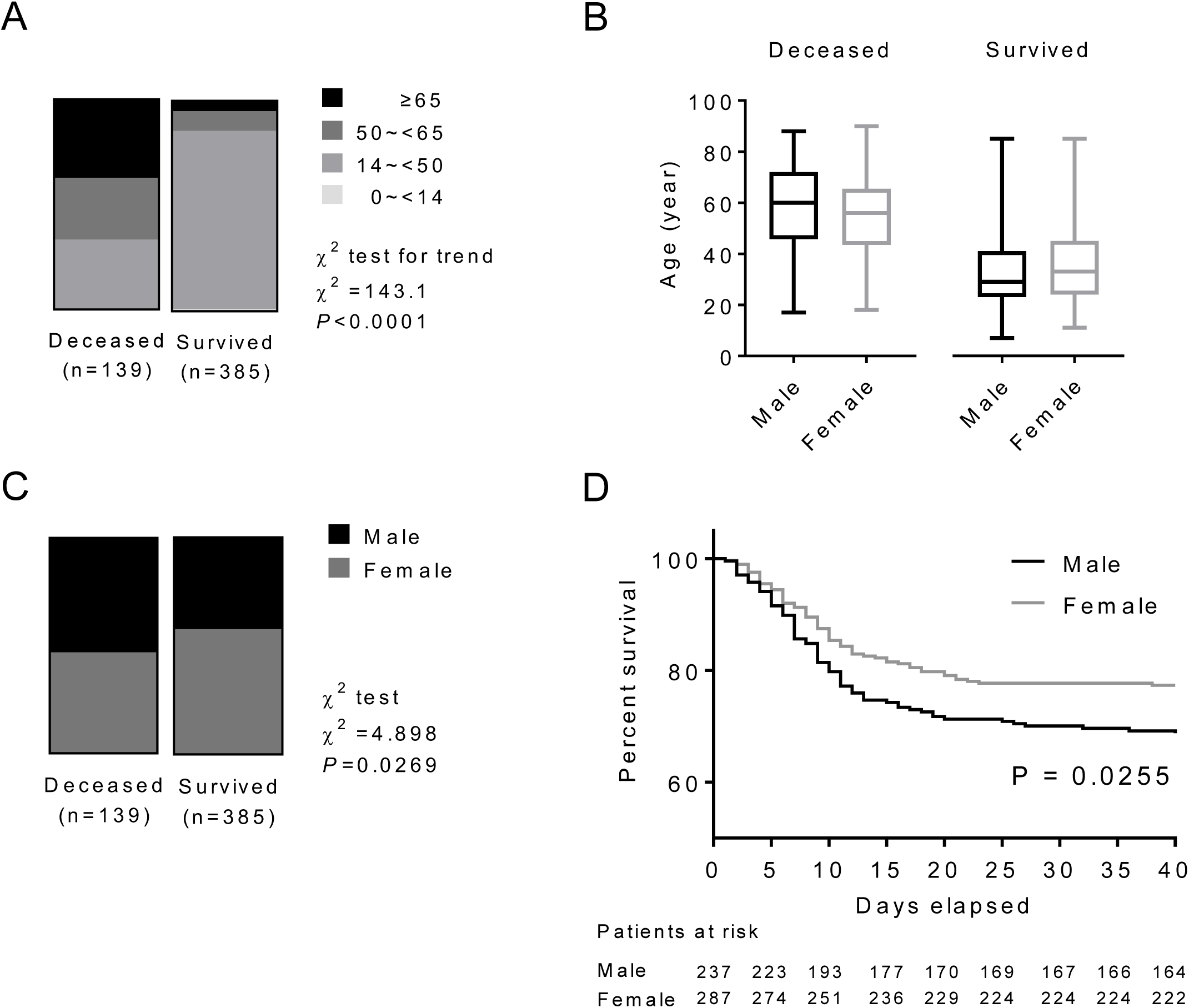
Role of age and gender in morbidity and mortality in patients with SARS. **(A)** The whole spectrum of age in deceased and survived patients with SARS. **(B)** Comparation of age between males and females in both deceased and survived patients with SARS. **(C)** Gender distribution in both deceased and survived patients with SARS. **(D)** Survival analysis comparing mortality rates between male and female patients with SARS.

## Discussion

Coronavirus is a large family of viruses that cause illness ranging from the common cold to severe pneumonia such as SARS^2^ and Middle East Respiratory Syndrome (MERS)^4^. SARS-CoV-2 was first identified in Wuhan city, China by the Chinese Center for Disease Control and Prevention (CDC)^5^. Both epidemiological^6,7^ and clinical^8,9^ features of patients with COVID-19 have recently been reported. However, few data on prognostic factors of COVID-19 have been reported.

Older age and the high number of comorbidities were associated with severity in patients with COVID-19^9-11^. Wang et al. reported that compared with patients not treated in the ICU (n=102) patients treated in the ICU (n=36) were older (median age, 66 years vs 51 years)^11^. Consistent with previous reports, older patients (≥65 years old), were more likely to have a severe type of COVID-19 in the case series patients in the present study. In the public data set, we also found that the percentage of older age (≥65 years) was much higher in the deceased patient than in the survived patients (83.8% in 37 deceased patients vs. 13.2% in 1019 survived patients). The results indicated that older age was associated with a high mortality in patients with COVID-19.

This is the first preliminary study investigating the gender role in morbidity and mortality of SARS-CoV-2 infection. It is suggested that COVID-19 is more likely to affect older males with comorbidities, and can result in severe and even fatal respiratory diseases such as ARDS^10^. Among the 425 patients with COVID-19, 56% were male, indicating males may be more susceptible to SARS-CoV-2 infection than females^6^. In 140 hospitalized patients with COVID-19, an approximately 1:1 ratio of male (50.7%) and female patients was found^10^. In four case series reports, 30 of 41(73%), 67 of 99 (68%), 25 of 51(49%) and 75 of 138 (54.3%) were men. In the present study, however, a similar susceptible to SARS-CoV-2 between males and females was observed in 1019 survived patients (50.0% males) collected from six cities and in a case series of 43 hospitalized patient (51.2% males).

Although the deceased patients were significantly older than the survived patients with COVID-19 in this study, ages were comparable between males and females in both the deceased and the survived patients. Consistent with this predominance, we also found that the proportion of male was higher in the deceased group than in the survived group in SARS patients. Survival analysis showed that men had a significantly higher mortality rate than women in SARS patients. Therefore, Gender is a risk factor for higher severity and mortality in patients with both COVID-19 and SARS independent of age and susceptibility.

In early 2003, we participated in the Beijing Municipal Medical Taskforce of SARS^12^. Here, we re-analyzed the data of a large population of 520 SARS patients including 135 deaths in Beijing, and summarized the experience and lessons for present use, because SARS-CoV-2 and SARS-CoV attack cells via the same receptor, ACE2^3^. We have previously reported that high protein expression of ACE2 receptor in specific organs correlated with specific organ failures indicated by corresponding clinical parameters in SARS patients^12,13^. Interestingly, the ACE2 gene is located on the X-chromosome and it has been shown that circulating ACE2 levels are higher in men than in women and in patients with diabetes or cardiovascular diseases^14^. Therefore, male patients may be more prone to die from SARS-CoV-2 because of the high expression of ACE2, though further research on the mechanism is needed.

### Limitations

This study has some limitations. First, for severity analysis only a case series of 43 patients with SARS-CoV-2 were included, because detailed patient information, particularly regarding clinical outcomes was unavailable in the public data set; Second, for mortality analysis only the first 37cases died of SARS-CoV-2 in Wuhan city were included. However, this is the first preliminary data investigating the gender role in morbidity and mortality in patients with SARS-CoV-2. More clinical and basic research regarding gender and other prognostic factors for individualized assessment and treatment is needed in the future.

## Conclusions

In conclusion, older age is a predominant risk factor for severity and morbidity in patients with both COVID-19 and SARS. While males and females have the same susceptibility to COVID-19, male patients may be more prone to dying independent of age.

## Data Availability

Address correspondence and reprint requests to Professor Jin-Kui YANG, Department of Medicine, Beijing Tongren Hospital, Capital Medical University, Beijing 100730, China
Tel: +86-10-58268445
Fax: +86-10-65288736
Cell: +86-13911167636

## Article Information

### Author Contributions

JMJ, PB, FG, WH, SL, FW, DMH SL and JKY collected the epidemiological and clinical data and processed statistical data. JKY drafted the manuscript. JMJ, SL and JKY revised the final manuscript. JKY is responsible for summarizing all epidemiological and clinical data.

## Acknowledgments

We thank all patients involved in the study.

